# Trends in Angiotensin Receptor Blocker Use Among those at Risk for COVID-19 Morbidity and Mortality in the United States

**DOI:** 10.1101/2020.07.24.20161851

**Authors:** Alexandra Perez, Robert Speth, Juan Saavedra

## Abstract

**Importance:** Assessment of the use of angiotensin receptor blockers (ARBs) in the United States provides insight into prescribing practices, and may inform guidelines, policy measures and research during the COVID-19 pandemic.

**Objective:** To evaluate trends in ARB use among adults in the United States who have preexisting conditions and sociodemographic risk factors that put them at a higher risk of SARS-CoV-2 infection and COVID-19-related complications and mortality.

**Design, setting and participants:** This study uses the nationally representative cross-sectional data from the 2005-2018 National Health and Nutrition Examination Survey (NHANES). Participants included 39,749 non-institutionalized U.S. civilian adults who were 20 years and older and those with the most common preexisting conditions and risk factors reported among patients with COVID-19.

**Main outcomes and measures:** Use of ARBs in the prior 30 days from survey interview.

**Results:** ARB use ranged from 7.4% [95% CI, 6.5%-8.4%] to 26.2% [95% CI, 19.4%-34.4%] among those with one or two metabolic, renal, respiratory, and/or cardiovascular diseases. Among individuals with the three most common preexisting conditions in patients with COVID-19 including hypertension, diabetes and obesity, ARB use was higher among the elderly, females, non-Hispanic whites, and those with health insurance coverage.

**Conclusions and relevance:** In this nationally representative survey, ARB use was found to be widespread, but unevenly distributed among individuals with conditions and sociodemographic risk factors that place them at a higher risk of COVID-19 morbidity and mortality.

**Key Points:** *Question:* What is the prevalence of angiotensin receptor blocker (ARB) use among individuals at higher risk of COVID-19-related complications?

*Findings:* In a cross-sectional study with data from 39,749 adult participants of the National Health and Nutrition Examination Survey, ARB use ranged between 7.4% and 26.2% among those with one or two respiratory, metabolic, renal and/or cardiovascular diseases. Significant disparities in ARB use were found in participants with preexisting conditions and sociodemographic factors that place them at a higher risk of COVID-19 morbidity and mortality.

*Meaning:* ARB use is widespread and disproportionate in the United States among people at higher risk of COVID-19 complications.

## INTRODUCTION

The COVID-19 pandemic affects not only the lung but multiple organs^1-7^ with high individual variability, in some cases with devastating consequences, leading to chronic disability or death.^8,9^ At the present time, no therapies have been devised to prevent or treat all cases of SARS-CoV-2 infection leading to COVID-19, however, a large number of therapeutic approaches have been proposed and several have been reported to have incremental benefits.^10-14^ Thus, it is imperative to identify available compounds that could be repurposed as beneficial, or at the very least palliative, to prevent or ameliorate the severity of the disease.

Some preexisting disorders such as hypertension, cardiovascular and kidney disease, diabetes, and obesity, are frequently comorbid with COVID-19.^15-24^ These disorders are characterized by enhanced activity of the Renin-Angiotensin System (RAS), in particular, with excessive stimulation of the Angiotensin (Ang) II AT1 receptor (AT1R) type.^25^ Increased AT1R activity leads to reduced systemic and pulmonary blood flow and oxygen levels, pathological inflammation, fibrosis, reduced sensitivity to insulin, alterations in innate and adaptive immunity, and coagulation defects.^26-28^ This explains why these preexisting conditions significantly increase the risk of poor outcome and death in COVID-19 patients.

It is not surprising that patients afflicted with these illnesses are frequently treated with medications such as Angiotensin Receptor Blockers (ARBs) that antagonize the pathological effects of AT1R hyperactivity.^**29-32**^ For these reasons, many patients afflicted with COVID-19 comorbidities and infected by SARS-CoV-2 have been previously treated with ARBs at the time of their diagnoses.

Although the use of ARBs for the treatment of hypertension, cardiovascular and metabolic disorders is widespread, data describing the precise prevalence of these medications in risk groups with comorbidities of COVID-19 is lacking. Such data are essential to address the question of the expected number of individuals treated with ARBs in the general population as well as at the time of SARS-CoV-2-infection. The objective of this study is to estimate ARB trends in use among adults in the United States who have preexisting conditions and sociodemographic risk factors that put them at a higher risk of SARS-CoV-2 infection and COVID-19-related complications and mortality. These data will be useful to determine, in future population studies, whether ARB use influences SARS-CoV-2 infection rates, the development of symptoms, the severity of the illness, the prognosis of COVID-19, and the process of recovery.

## METHODS

### Data Source and Study Population

The National Health and Nutrition Examination Survey (NHANES) is a nationally representative cross-sectional survey of the non-institutionalized civilian population in the United States.^33^ This survey consists of questionnaire, physical examination and laboratory components that includes self-reported data of medical history, prescription medication use, and body measures among others. This study used publicly available, deidentified data and did not require institutional review board approval (45 CFR§46.102(f)).

We used data from adults 20 years of age or older and evaluated ARB use within the most prevalent (5% or higher) preexisting conditions reported in the COVID-19 population.^34^ For this study, data were used from the 7 most recent NHANES biannual cycles being 2005 through 2018.

### Assessment of Angiotensin Receptor Blocker Use

The prescription medications survey provides personal interview data on the use of prescription medications during a 30-day period prior to the participant’s interview date. All prescription drugs are entered upon interview by generic name. ARB use was defined as use of any angiotensin receptor blocker (losartan, candesartan, olmesartan, azilsartan, eprosartan, irbesartan, telmisartan, and valsartan) either as a single agent or in combination with other drugs.

### Statistical Analysis

This study assessed the prevalence of ARBs in the general adult population and within populations of hypertension, diabetes, coronary heart disease, stroke, kidney disease, congestive heart failure, obesity, asthma, and chronic obstructive pulmonary disease (COPD) as shown in Table 1. ARB use was also explored within select populations with 2 concomitant metabolic, renal and cardiovascular diseases and data are presented in eTable 1 in the Supplement. ARB use within NHANES biannual cycle was also assessed in the general adult population and within the previously specified conditions from 2005-2018 (Figure 1 and eTable 2 in the Supplement). Furthermore, the association of ARB use and commonly reported sociodemographic risk factors in those diagnosed with COVID-19 including age, gender, race/ethnicity, household income and educational levels, health and prescription insurance coverage and body mass index were explored within populations with hypertension, diabetes and obesity (defined as ≥30 kg/m^2^) and are shown in Tables 2-4. These conditions are the three most prevalent comorbidities (30% or higher) among those with COVID-19.^34^ Similar analyses were conducted within populations with coronary heart disease, stroke, kidney disease and congestive heart failure and are presented in eTable 3 and eTable 4 in the Supplement. Sociodemographic risk factor heterogeneity was assessed by stratification of age (≤44, 45-64 and ≥65 years), sex (male/female), race/ethnicity (non-Hispanic white, non-Hispanic black, Hispanic, and other race), income (≥$20,000, <$20,000), education (high school or lower, some college or higher), health insurance coverage (yes/no), prescription insurance coverage (yes/no) and body mass index (≤24.99 kg/m^2^, 25-29.99 kg/m^2^, 30-39.99 kg/m^2^, and ≥40 kg/m^2^). Prevalent ARB use was calculated within unique risk factor subgroups (e.g., within participants age ≤ 44) and distribution across subgroups (e.g., across all age categories). Survey-weighted binomial logistic regression evaluated the association of ARB use, as the dependent variable, and biannual cycle, sociodemographic risk factors and body mass index categories were independent parameters. ARB use prevalence estimates were adjusted for oversampling of older adults, low-income individuals, and racial/ethnic minorities for their unequal sampling probability and nonresponse. Statistical significance was assessed at the 2-sided α=0.05 level. All analyses were conducted using IBM SPSS version 26.0.

**Figure 1.**
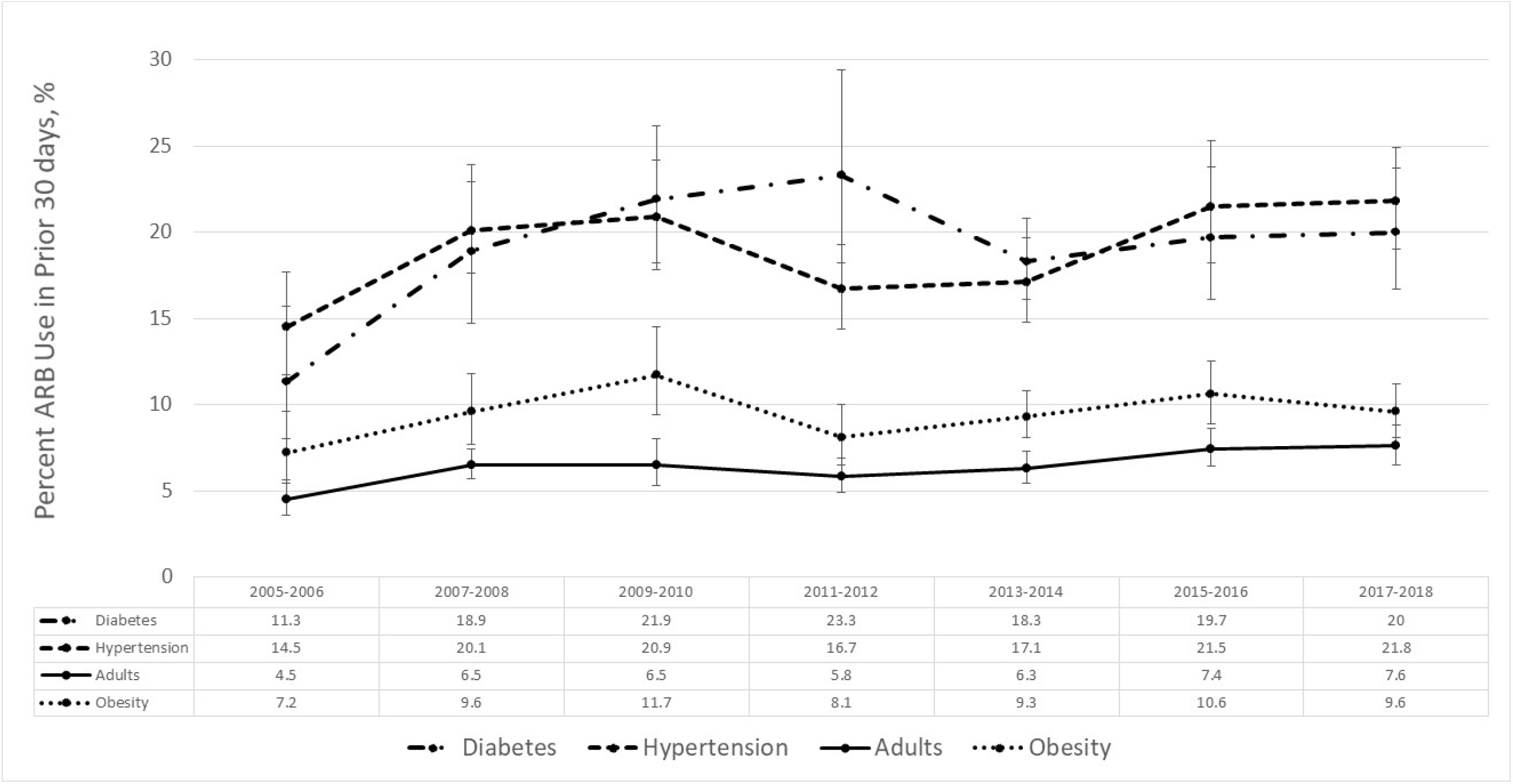
Trends in Angiotensin Receptor Blocker Use Across NHANES Biannual Cycles 2005-2018 Among Adults and by Diabetes, Hypertension and Obesity Diagnoses.

**Figure 1.**
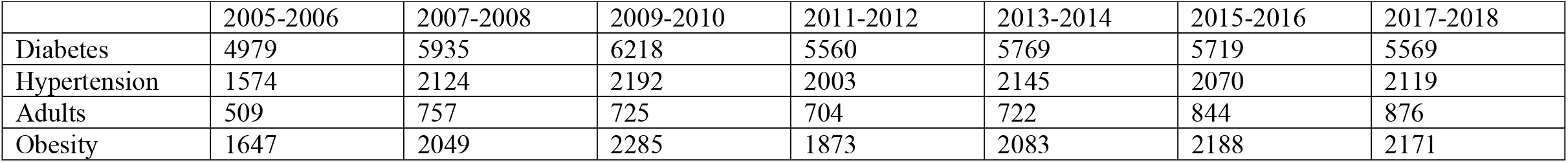
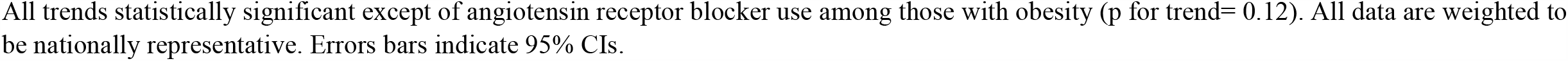
Number of Adult Participants Across NHANES Biannual Cycles 2005-2018 Overall and by Diabetes, Hypertension and Obesity Diagnoses.

**Table 1.**
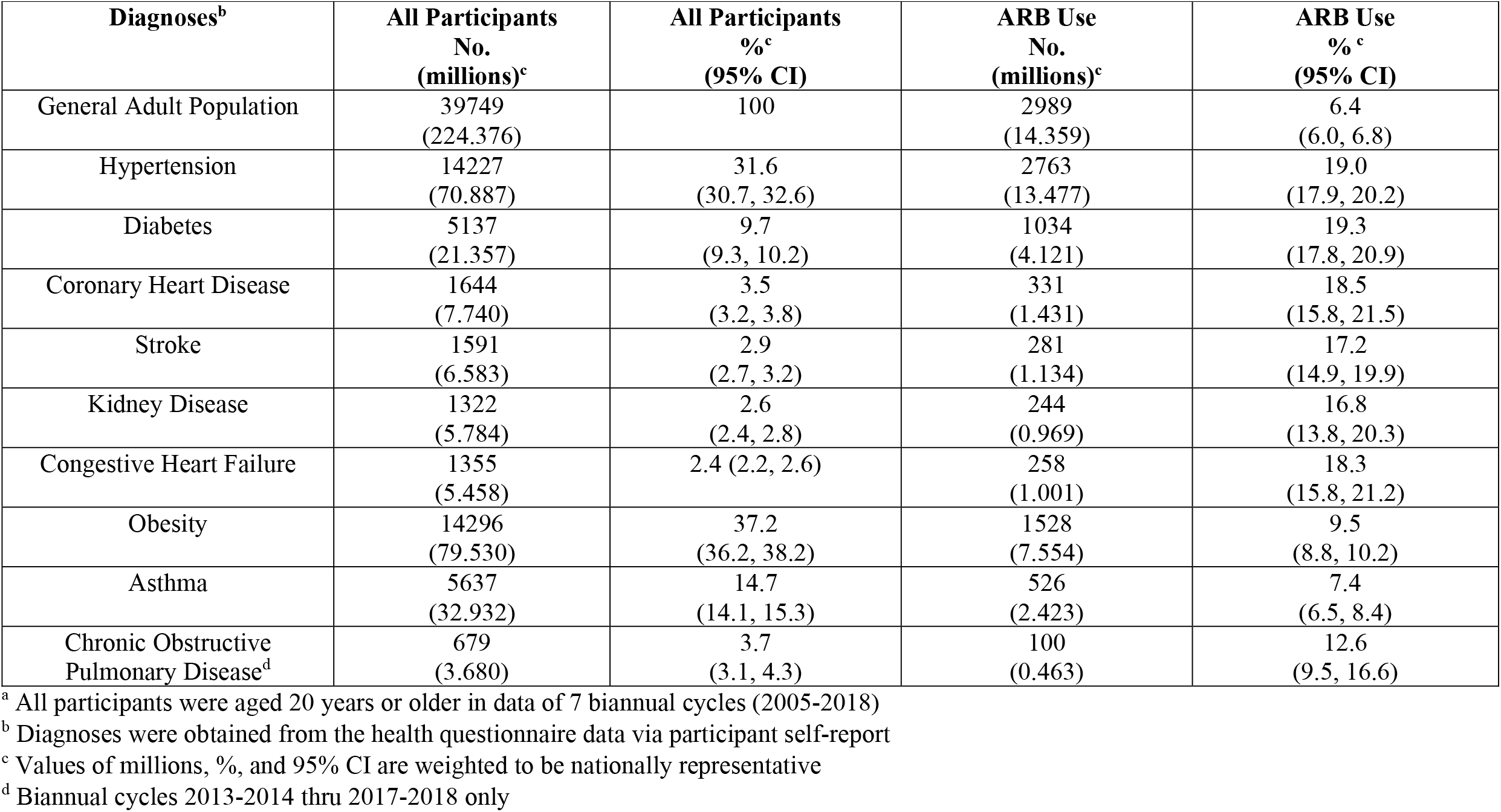
Prevalence of ARB use Among US Adults Overall and Those with Select Conditions^a^.

**Table 2.**
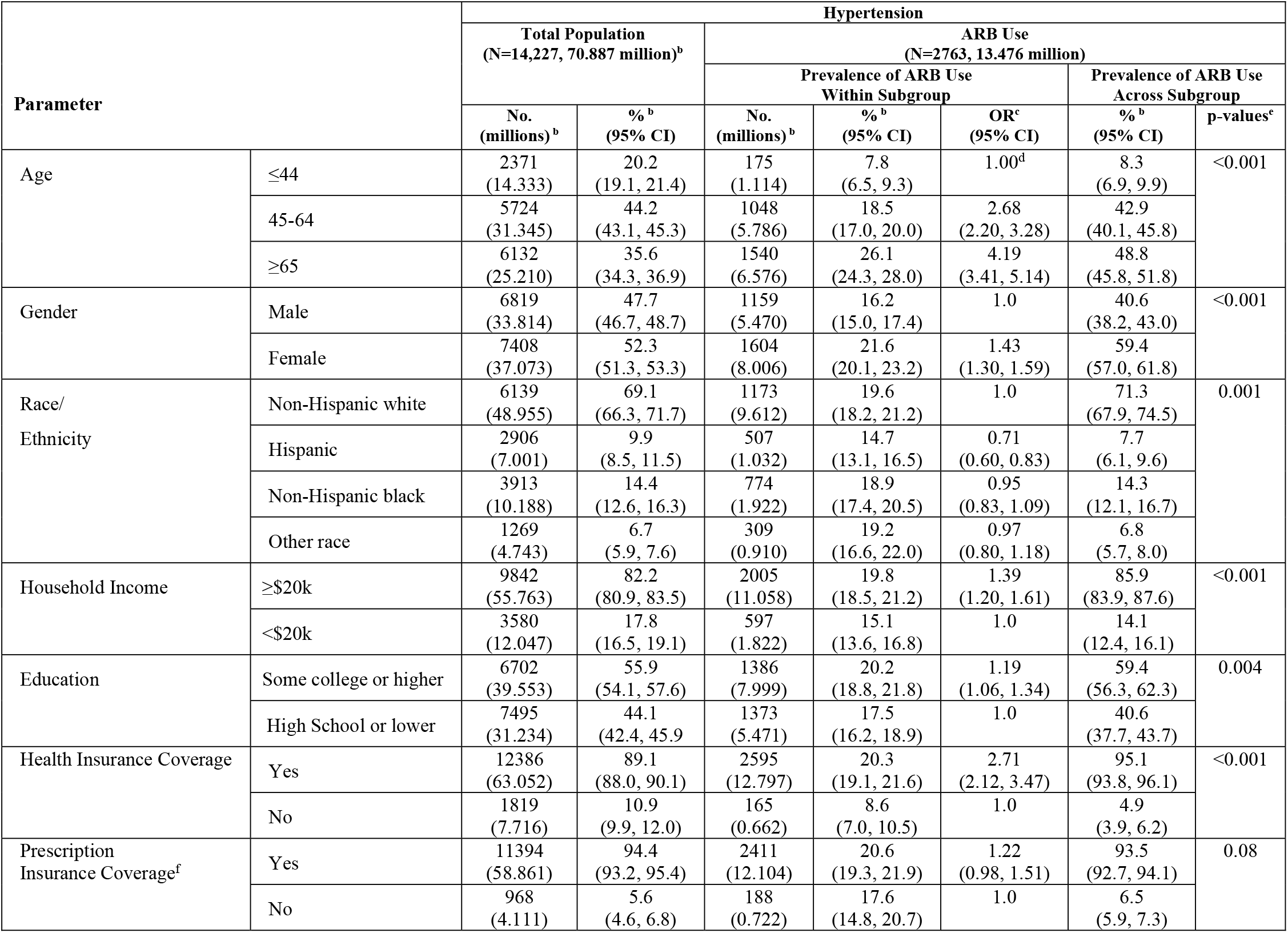
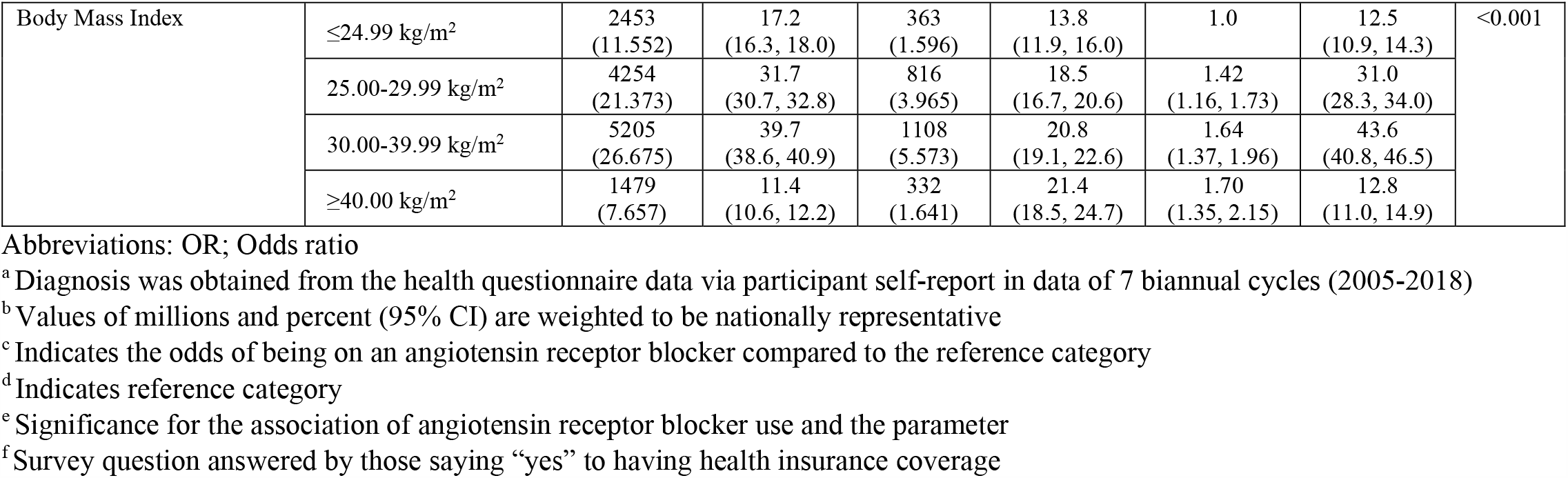
Prevalence of Angiotensin Receptor Blocker Use in the Prior 30 days Among US Adults with Hypertension by Sociodemographic Characteristics and Body Mass Index^a^.

**Table 3.**
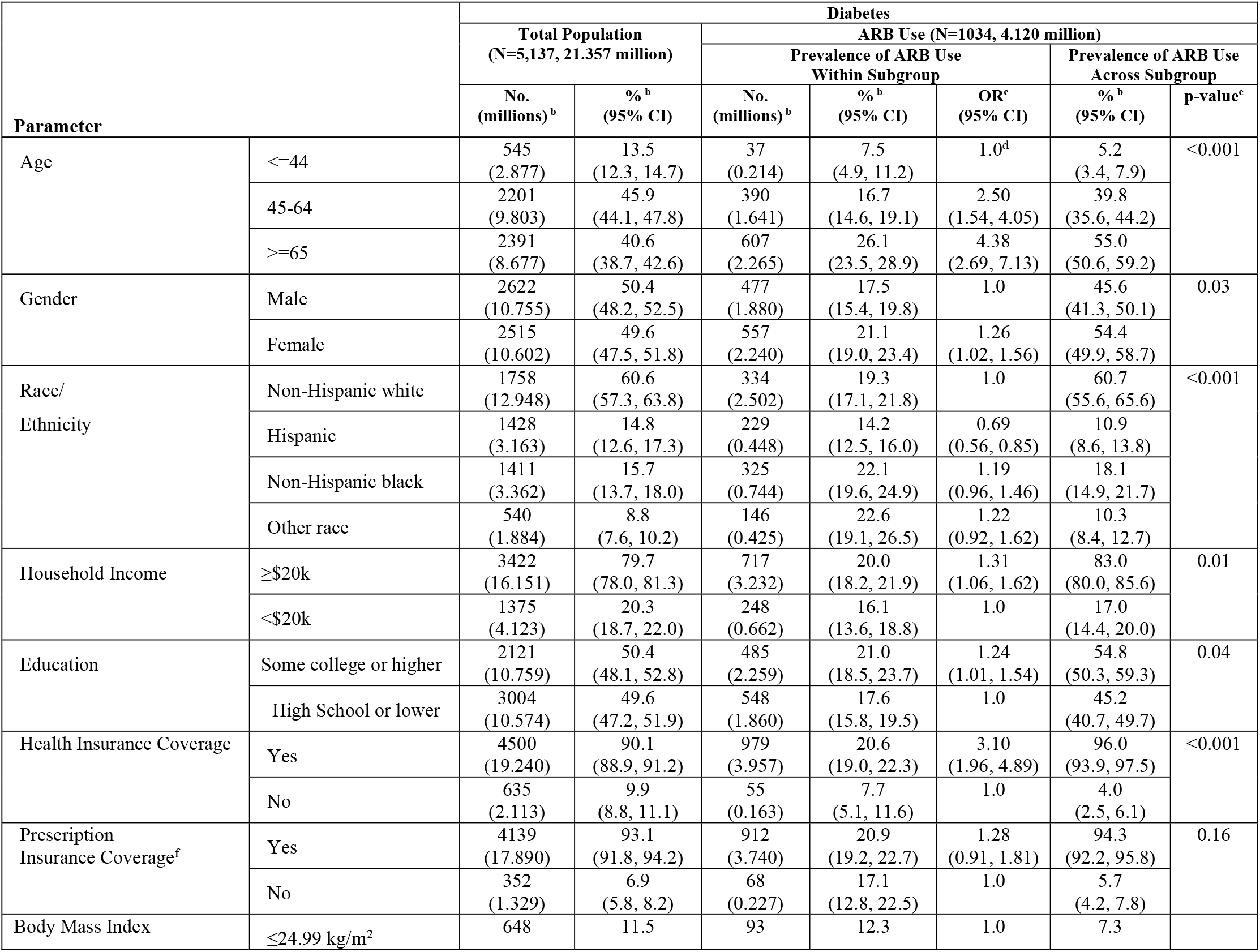
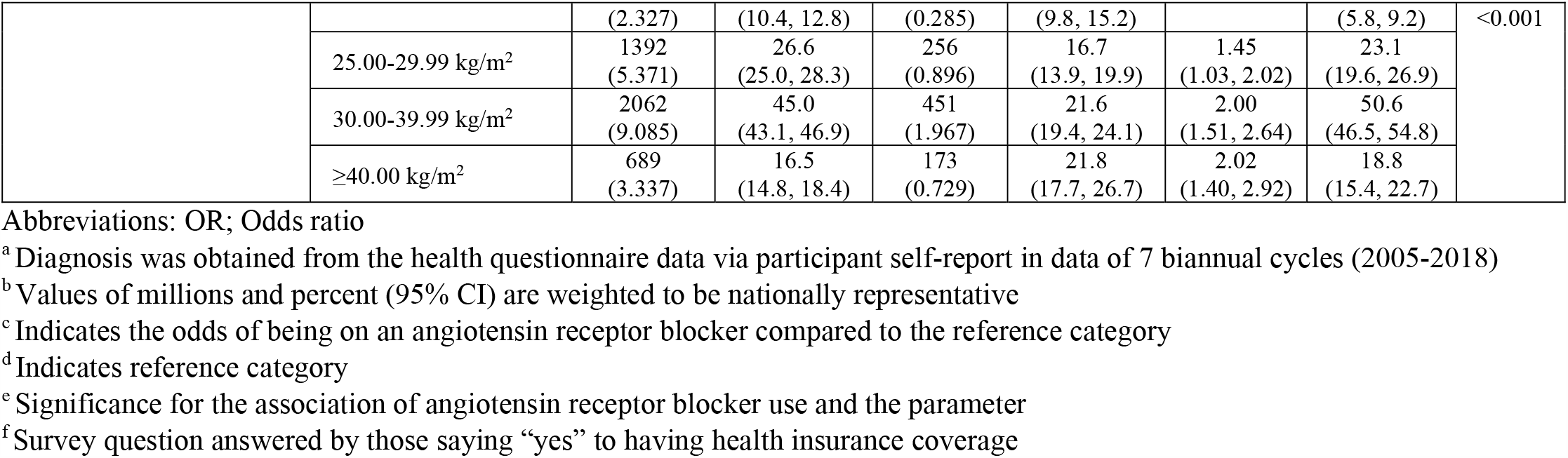
Prevalence of Angiotensin Receptor Blocker Use in the Prior 30 days Among US Adults with Diabetes by Social Determinants of Health and Body Mass Index^a^.

**Table 4.**
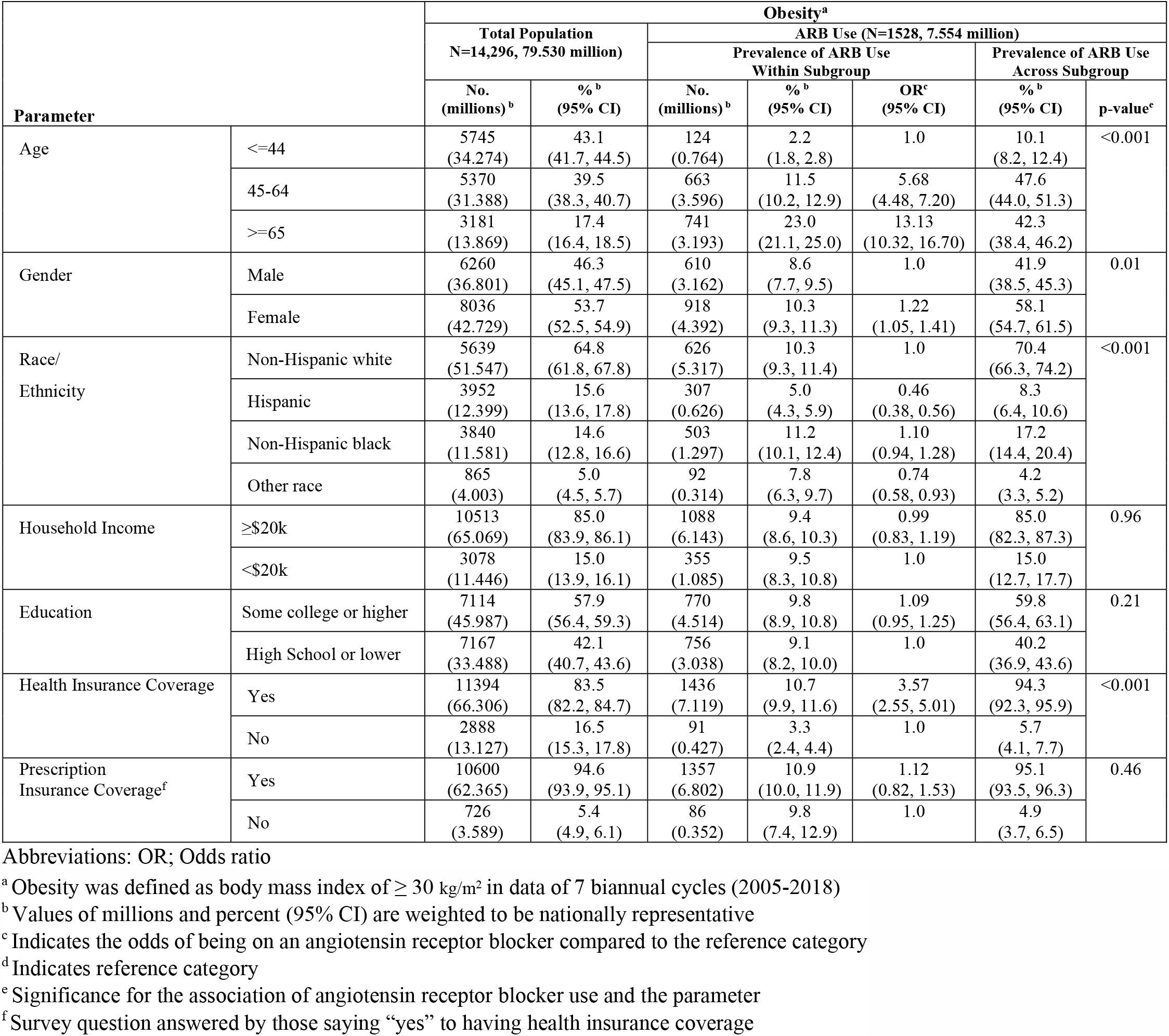
Prevalence of Angiotensin Receptor Blocker Use in the Prior 30 days Among US Adults with Obesity by Social Determinants of Health and Body Mass Index^a^.

| Parameter |  | Obesity<sup>a</sup> |  |  |  |  |  |  |
| --- | --- | --- | --- | --- | --- | --- | --- | --- |
|  |  | Total Population<br>N=14,296, 79.530 million) |  | ARB Use (N=1528, 7.554 million) |  |  |  |  |
|  |  |  |  | Prevalence of ARB Use<br>Within Subgroup |  |  | Prevalence of ARB Use<br>Across Subgroup | p-value<sup>e</sup> |
|  |  | No.<br>(millions)<sup>b</sup> | %<sup>b</sup><br>(95% CI) | No.<br>(millions)<sup>b</sup> | %<sup>b</sup><br>(95% CI) | OR<sup>c</sup><br>(95% CI) | %<sup>b</sup><br>(95% CI) |  |
| Age | <=44 | 5745<br>(34.274) | 43.1<br>(41.7, 44.5) | 124<br>(0.764) | 2.2<br>(1.8, 2.8) | 1.0 | 10.1<br>(8.2, 12.4) | <0.001 |
|  | 45-64 | 5370<br>(31.388) | 39.5<br>(38.3, 40.7) | 663<br>(3.596) | 11.5<br>(10.2, 12.9) | 5.68<br>(4.48, 7.20) | 47.6<br>(44.0, 51.3) |  |
|  | >=65 | 3181<br>(13.869) | 17.4<br>(16.4, 18.5) | 741<br>(3.193) | 23.0<br>(21.1, 25.0) | 13.13<br>(10.32, 16.70) | 42.3<br>(38.4, 46.2) |  |
| Gender | Male | 6260<br>(36.801) | 46.3<br>(45.1, 47.5) | 610<br>(3.162) | 8.6<br>(7.7, 9.5) | 1.0 | 41.9<br>(38.5, 45.3) | 0.01 |
|  | Female | 8036<br>(42.729) | 53.7<br>(52.5, 54.9) | 918<br>(4.392) | 10.3<br>(9.3, 11.3) | 1.22<br>(1.05, 1.41) | 58.1<br>(54.7, 61.5) |  |
| Race/<br>Ethnicity | Non-Hispanic white | 5639<br>(51.547) | 64.8<br>(61.8, 67.8) | 626<br>(5.317) | 10.3<br>(9.3, 11.4) | 1.0 | 70.4<br>(66.3, 74.2) | <0.001 |
|  | Hispanic | 3952<br>(12.399) | 15.6<br>(13.6, 17.8) | 307<br>(0.626) | 5.0<br>(4.3, 5.9) | 0.46<br>(0.38, 0.56) | 8.3<br>(6.4, 10.6) |  |
|  | Non-Hispanic black | 3840<br>(11.581) | 14.6<br>(12.8, 16.6) | 503<br>(1.297) | 11.2<br>(10.1, 12.4) | 1.10<br>(0.94, 1.28) | 17.2<br>(14.4, 20.4) |  |
|  | Other race | 865<br>(4.003) | 5.0<br>(4.5, 5.7) | 92<br>(0.314) | 7.8<br>(6.3, 9.7) | 0.74<br>(0.58, 0.93) | 4.2<br>(3.3, 5.2) |  |
| Household Income | ≥\$20k | 10513<br>(65.069) | 85.0<br>(83.9, 86.1) | 1088<br>(6.143) | 9.4<br>(8.6, 10.3) | 0.99<br>(0.83, 1.19) | 85.0<br>(82.3, 87.3) | 0.96 |
| | <\$20k | 3078<br>(11.446) | 15.0<br>(13.9, 16.1) | 355<br>(1.085) | 9.5<br>(8.3, 10.8) | 1.0 | 15.0<br>(12.7, 17.7) | |
| Education | Some college or higher | 7114<br>(45.987) | 57.9<br>(56.4, 59.3) | 770<br>(4.514) | 9.8<br>(8.9, 10.8) | 1.09<br>(0.95, 1.25) | 59.8<br>(56.4, 63.1) | 0.21 |
|  | High School or lower | 7167<br>(33.488) | 42.1<br>(40.7, 43.6) | 756<br>(3.038) | 9.1<br>(8.2, 10.0) | 1.0 | 40.2<br>(36.9, 43.6) |  |
| Health Insurance Coverage | Yes | 11394<br>(66.306) | 83.5<br>(82.2, 84.7) | 1436<br>(7.119) | 10.7<br>(9.9, 11.6) | 3.57<br>(2.55, 5.01) | 94.3<br>(92.3, 95.9) | <0.001 |
|  | No | 2888<br>(13.127) | 16.5<br>(15.3, 17.8) | 91<br>(0.427) | 3.3<br>(2.4, 4.4) | 1.0 | 5.7<br>(4.1, 7.7) |  |
| Prescription<br>Insurance Coverage<sup>f</sup> | Yes | 10600<br>(62.365) | 94.6<br>(93.9, 95.1) | 1357<br>(6.802) | 10.9<br>(10.0, 11.9) | 1.12<br>(0.82, 1.53) | 95.1<br>(93.5, 96.3) | 0.46 |
|  | No | 726<br>(3.589) | 5.4<br>(4.9, 6.1) | 86<br>(0.352) | 9.8<br>(7.4, 12.9) | 1.0 | 4.9<br>(3.7, 6.5) |  |

Abbreviations: OR; Odds ratio
- <sup>a</sup> Obesity was defined as body mass index of $\geq 30$ kg/m<sup>2</sup> in data of 7 biannual cycles (2005-2018) - <sup>b</sup> Values of millions and percent (95% CI) are weighted to be nationally representative - <sup>c</sup> Indicates the odds of being on an angiotensin receptor blocker compared to the reference category - <sup>d</sup> Indicates reference category - <sup>e</sup> Significance for the association of angiotensin receptor blocker use and the parameter - <sup>f</sup> Survey question answered by those saying “yes” to having health insurance coverage

## RESULTS

Data from 39,749 participant adults aged 20 years and older were used in this study and the sample size for each biannual NHANES cycle ranged from 4979 to 6218. Sample size for populations with hypertension, diabetes and obesity are presented in Tables 2-4. Response rates in NHANES cycles ranged between 71% and 94.3%.^35^ Table 1 shows the national count and percent of US adults reporting use of an ARB in the general adult population and within populations with select metabolic, respiratory, renal and cardiovascular diseases. 6.4% percent (or 14.4 million people) in the general adult population, regardless of comorbidities, in the US were estimated to be taking ARBs. In the single condition population analysis in Table 1, ARB use was at its lowest among those with asthma (7.4% [95% CI, 6.5%-8.4%, 2.4 million people]) and at its highest among those with diabetes (19.3% [95% CI, 17.8%-20.9%, 4.1 million people]). ARB use was more prevalent within populations with 2 co-existing chronic conditions. As shown in eTable 1 in the Supplement, ARB use prevalence was at its lowest in those with co-existing kidney disease and congestive heart failure (18.7% [95% CI, 12.8%-26.5%, 175,000 people] and at its highest among those with both coronary heart disease and kidney disease (26.2% [95% CI, 19.4%-34.4%, 197,000 people]).

Figure 1 reports a significant increase in the prevalence of ARB use from 2005 to 2018 in the general adult population (4.5% [95% CI, 3.6%-5.6%] in 2005-2006 and 7.6% [95% CI, 6.5%-8.8%] in 2017-2018, p for trend=0.007). The same trend was observed among those with hypertension (14.5% [95% CI, 11.7%-17.7%] in 2005-2006 and 21.8% [95% CI, 19.0%-24.9%] in 2017-2018, p for trend =0.005), and diabetes (11.3% [95% CI, 8.0%-15.7%] in 2005-2006 and 20.0% [95% CI, 16.7%-23.7%] in 2017-2018,p for trend =0.02) but not in those with obesity (7.2% [95% CI, 5.4%-9.6%] in 2005-2006 and 9.6% [95% CI, 8.1%-11.2%] in 2017-2018, p for trend =0.12). ARB use among participants with coronary heart disease, stroke, kidney disease, and congestive heart failure did not change across biannual cycles and data are shown in eTable 2 in the Supplement.

Tables 2-4 show the prevalence of ARB use by select sociodemographic risk factors and body mass index categories among those with hypertension, diabetes, and obesity. In these three populations, there was a significant association between ARB use and age groups (p<0.001 for all), gender (p<0.03 for all), race/ethnicity (p<0.001 for all), and health insurance coverage (p<0.001 for all). In those with hypertension and diabetes, a significant association of ARB use and household income (p<0.01 for both), educational level (p<0.04 for both) and body mass index category (p<0.0001 for both) was observed. Odds ratios in Tables 2-4 indicate that ARB use in those with hypertension, diabetes and obesity increased with age, was higher among females than males, non-Hispanic whites compared to minorities and in those with health insurance coverage. In those with hypertension and diabetes, ARB use was higher among those with household income ≥$20,000, some college education or higher, and higher body mass index level. Prescription insurance coverage was not associated with ARB use in any of the populations evaluated. eTables 3 and eTable 4 in the Supplement show the prevalence of ARB use among participants with coronary heart disease, stroke, kidney disease, and congestive heart failure and show similar trends in ARB use within sociodemographic risk factors.

## DISCUSSION

Our results demonstrate that ARB use is prevalent for conditions that increase the risk of SARS-CoV-2 infection and increase COVID-19 morbidity and mortality. ARB use in the United States in the general adult population is 6.4% and ranging between 7.4% and 26.2% among those with one or two metabolic, respiratory, renal and cardiovascular diseases prevalent in the COVID-19 population. Over time, ARB use has increased in the general population and among those with hypertension and diabetes but not in the other conditions evaluated. Overall, ARB use was most prevalent with increasing age, among females, non-Hispanic whites, those with health insurance coverage, and higher income, educational level and body mass index categories in many of the populations evaluated including hypertension, diabetes, obesity and other cardiovascular diseases.

Along with other antihypertensive agents, ARBs are currently indicated to be initial treatment for hypertension among non-Blacks and as secondary treatment for Blacks.^36^ However, ARBs or ACE inhibitors are recommended for treatment of hypertension in all patients with diabetes.^36^ ARBs are very well tolerated, including in the elderly, increasingly available in the therapeutic arsenal as generic drugs, and with significant beneficial properties beyond their effect on blood pressure, such as potently reducing inflammation and fibrosis and mitigating the adverse effects of metabolic syndrome.^37-41^ As a consequence, there is increasing interest to consider ARB treatment for a larger number of disorders beyond hypertension. There are 1,724 clinical studies presently testing the use of ARBs not only for cardiovascular disease, but for many other clinical conditions.^42^ For these reasons it is expected that a large and increasing number of patients will be medicated with ARBs at the time of their COVID-19 diagnosis.

Initial concerns for continued use of ARBs in COVID-19 patients have been rebutted, with the consensus now being that ARBs should not be withdrawn when prescribed for comorbidities, since they do not adversely influence COVID-19 prognosis and progression.^37,40,43-47^ Moreover, it is possible that treatment with ARBs might actually be beneficial, since these compounds protect lung function, reducing pneumonia-related deaths associated with hyper-inflammation.^48-52^ Preliminary reports now suggest that ARBs may reduce COVID-19 disease severity.^53-55^ A large-scale observational study of hospitalized COVID-19 patients with preexisting hypertension demonstrated a profound reduction in mortality in patients taking ARBs compared to other antihypertensive drugs.^55^

Nevertheless, it is most important to clearly determine whether ARBs could be beneficial to ameliorate or delay COVID-19 progress, reduce mortality and improve health during recovery, and the effects of ARBs on COVID-19 patients are under intense scrutiny. To this date (07/17/2020), there are 45 ongoing prospective clinical trials registered with clinicaltrials.gov ^42^ to determine the precise impact of ARB use in COVID-19 patients.

Our data also demonstrate that while ARBs are frequently prescribed for patients with COVID-19 comorbidities, ARB use is not uniform within the US population. The incidence of COVID-19 is higher among individuals with lower socioeconomic status who are subject to increased exposure to SARS-CoV-2 by virtue of their need to continue working in environments prone to contagion, as well as living in a crowded home environment. COVID-19 incidence is also higher in elderly, males, racial/ethnic minority (i.e., Hispanics, Blacks and Native Americans) populations, as well as those patients previously affected by hypertension, diabetes, obesity, cardiovascular and kidney disease; considered to be risk factors and co-morbidities.^15-24^ In this study, we found ARB use to be lowest in these sociodemographic subgroups.

For these reasons, determination of ARB benefit should be tempered by the consideration that those individuals treated with ARBs may have better overall physical health and less intense exposure to the coronavirus, confounding factors to consider when reaching final conclusions on ARB benefits.

Comparison of incidence and outcomes of COVID-19 for ARB use compared with other RAS inhibitors such as ACE inhibitors, and other classes of antihypertensive drugs are under way.^42^

Our study has several limitations: 1) since survey participants were non-institutionalized civilians, the study results may not be extrapolated to those living in nursing homes or incarcerated individuals where COVID-19 has been prevalent and should only be generalized to the community-dwelling US adult population; 2) diagnoses of the comorbidities and medication use presented here are based on self-report and may be subject to error due to recall bias; and 3) this study does not address the use of single ARB therapy compared with the use of ARBs combined with additional antihypertensive drugs or compliance to treatment. Evaluation of the use of ARBs among the highly affected Native American population was not possible in this study.

In conclusion, our report will inform both clinical practice and research, and will be useful in prioritizing future studies on whether ARB use influences SARS-CoV-2 infection rates, the development of symptoms, the severity of the illness, the prognosis of COVID-19, and the process of recovery in the subpopulations evaluated here.

## Data Availability

Data may be shared upon request at the discretion of authors.

## Supplement

**eTable 1.**
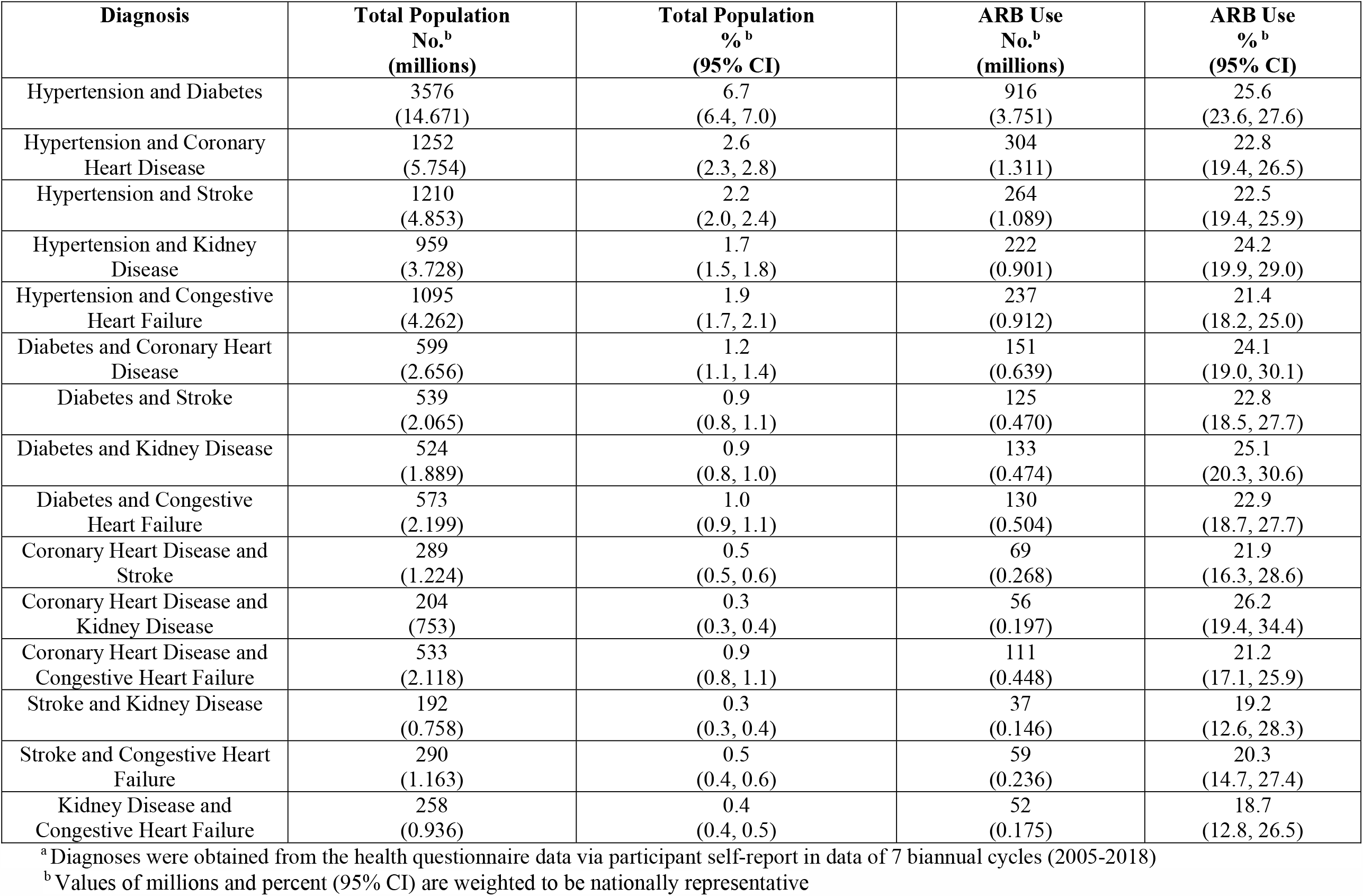
Prevalence of ARB Use in Prior 30 days Among Adults in the United States who Reported Being Diagnosed with Two Select Metabolic, Renal and Cardiovascular Diseases^a^.

**eTable 2.**
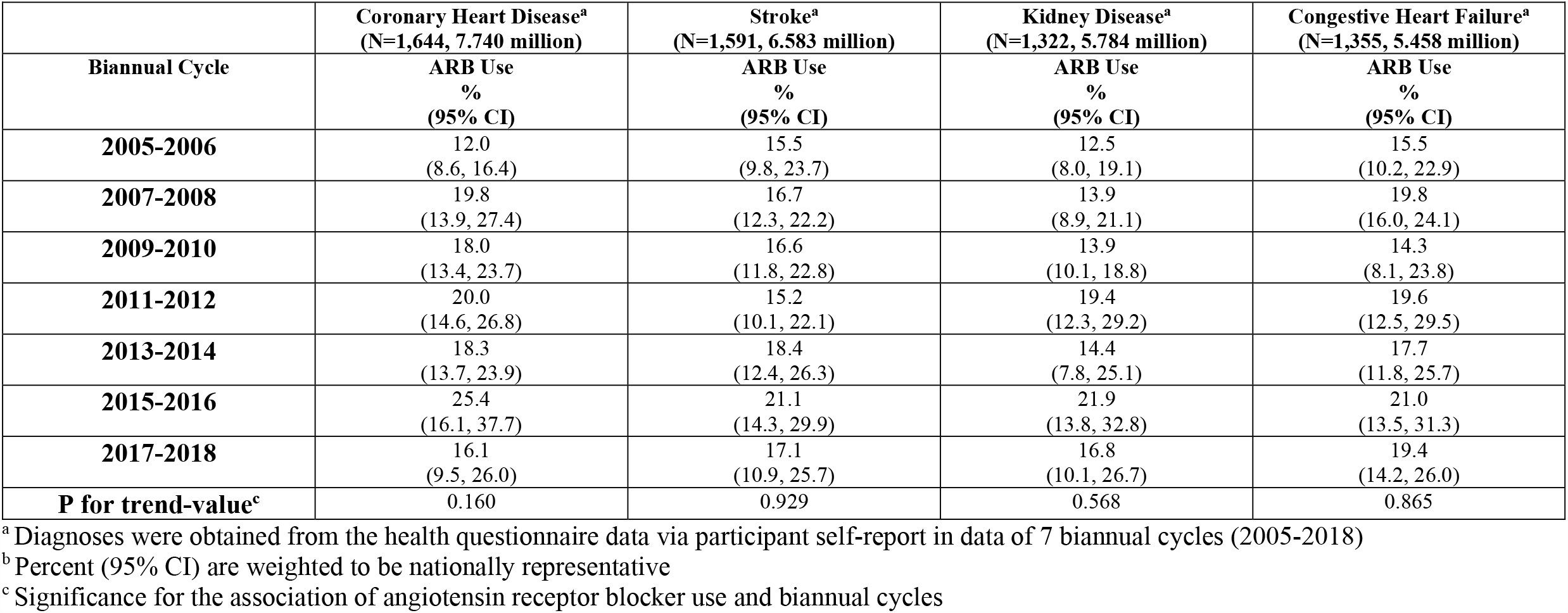
Trends in Angiotensin Receptor Blocker Use in the US in the Prior 30 Days across NHANES biannual cycles 2005-2018 Among Adults with Coronary Heart Disease, Stroke, Kidney Disease and Congestive Heart Failure.

**eTable 3.**
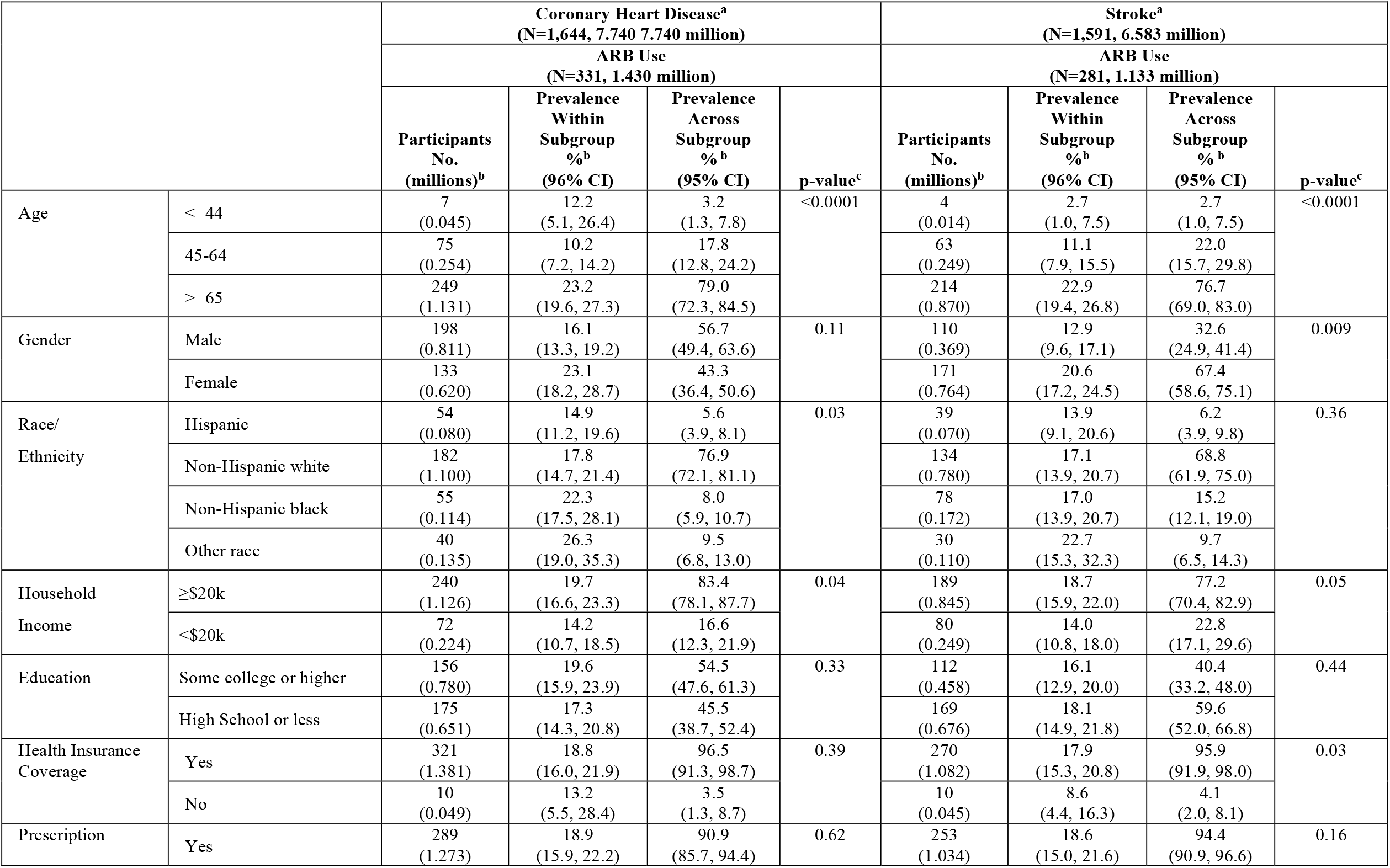
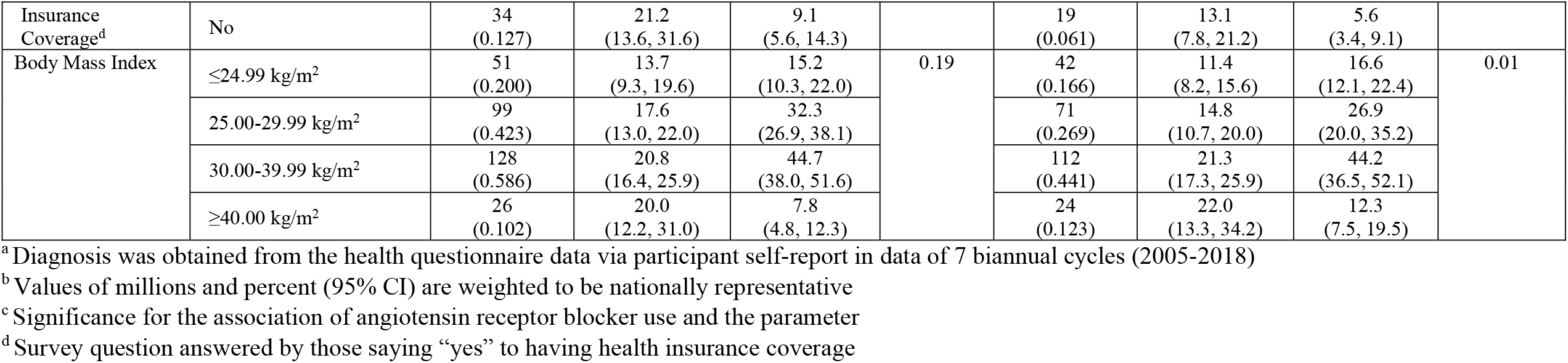
Prevalence of Angiotensin Receptor Blocker Use in the Prior 30 days Among US Adults with Coronary Heart Disease and Stroke by Social Determinants of Health and Body Mass Index.

**eTable 4.**
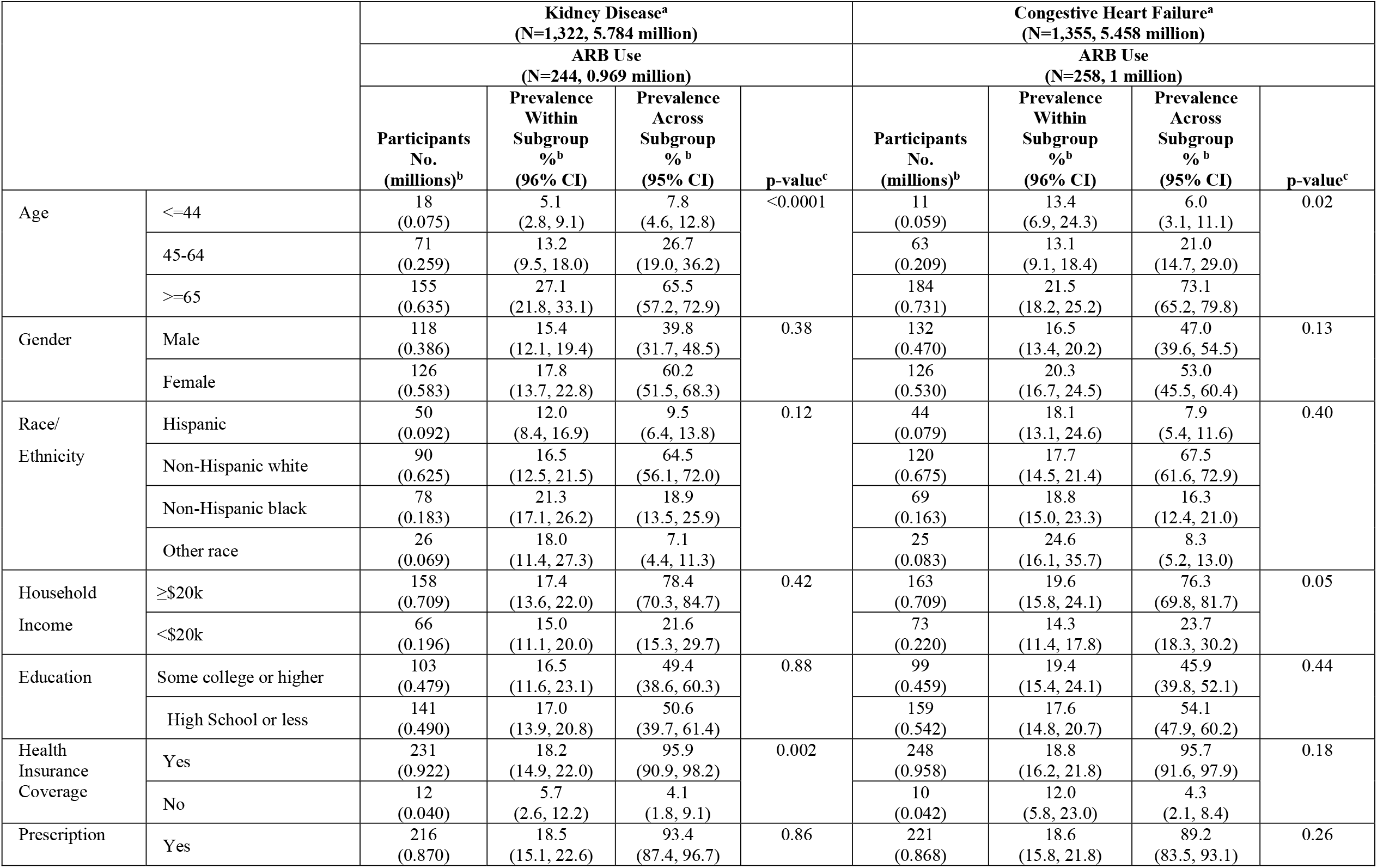
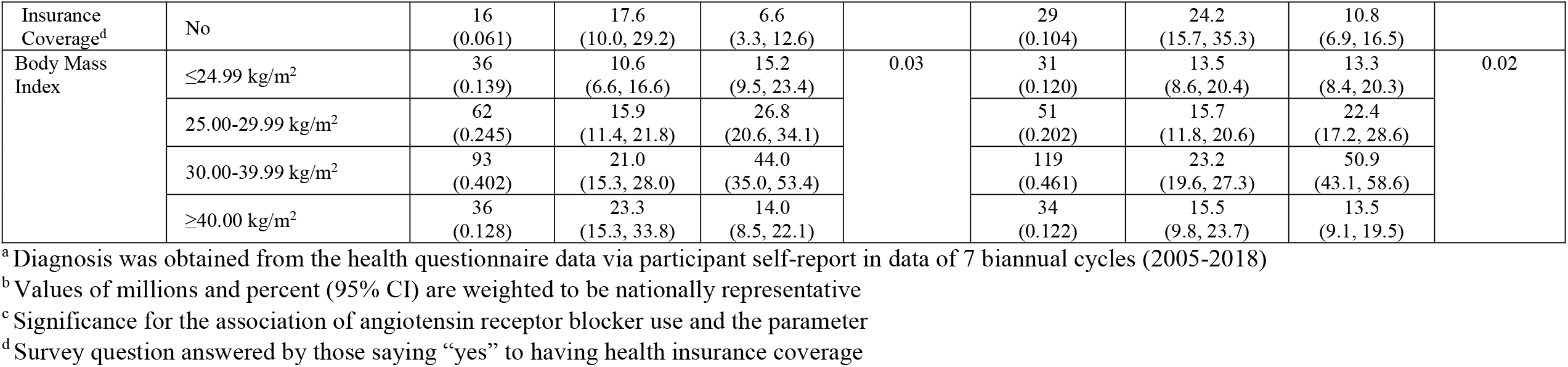
Prevalence of Angiotensin Receptor Blocker Use in the Prior 30 days Among US Adults with Kidney Disease and Congestive Heart Failure by Social Determinants of Health and Body Mass Index.

